# Cancer risk perception and knowledge and physician communication behaviors: specific influences on cervical cancer and colorectal cancer screening in women 50-65 years old

**DOI:** 10.1101/2021.05.04.21256488

**Authors:** Diane M Harper, Madiha Tariq, Asraa Alhawli, Nadia Syed, Minal R Patel, Ken Resnicow

## Abstract

**Background:** Women 50-65 years old have the lowest cervical and colorectal cancer (CRC) screening rates amongst ages recommended for screening. The primary aim of this work is to determine how cancer risk perceptions and provider communication behaviors, in addition to known demographic factors, influence the uptake of both cervical and CRC screening or a single screen among women in southeast Michigan.

**Methods:** 14 health services and communication behavior questions were adapted from the Health Information National Trends Survey (HINTS) and administered to a multiethnic sample of adults in southeast Michigan. The outcome variable was self-reported up to date cervical and/or CRC screening as defined by the United States Preventive Services Task Force (USPSTF). Demographic and cancer risk/communication behavior responses of the four screening status categories (both tests, one test, no tests) were analyzed with multinomial regression for all comparisons.

**Results:** Of the 394 respondents, 54% were up to date for both cervical and CRC screening, 21% were up to date with only cervical cancer screening and 12% were up to date for only CRC screening. Of the 14 risk perception and communication behaviors only, Did your primary care physician (PCP) involve you in the decisions about your health care as much as you wanted? was significantly associated with women having both screens compared to only cervical cancer screening (aOR 1.67 (95% CI: 1.08, 2.57). The control variables included in the model, also significant, were MENA and Black races compared to white women and age.

**Conclusions:** Screening behavior for both cancers is different than a single screen and associated with the woman’s perception of the physician involving her in her care as much as she wanted. In addition, educational programs are needed for MENA women.

## Introduction

Women 50-65 years of age have the lowest cervical cancer screening rate among those 21-65 years old, as reported in three national surveillance databases: Behavioral Risk Factor Surveillance System (BRFSS), Health Information National Trends Survey (HINTS) and Health Center Patient Survey (HCPS) (**Harper DM 2020)**.^**1**^ Likewise, women 50-65 years old also have the lowest colorectal cancer (CRC) screening rates among those 50-75 years old (**ACS2020**).^2^ Only half of the women in this age group are up to date for both screens while, 25% have cervical only, 12% CRC only and 12% have neither screen (**Harper DM 2021**).^3^ As we have previously reported, women more likely to be up to date for only cervical cancer screening compared to both cancer screens are younger, and are MENA compared to white race (**Harper DM 2021**).^3^

Racial disparities in cervical cancer screening are prominent among Middle Easter/North African (MENA) women in the US (**Harper DM 2021**)^4^. MENA women between 50-65 years old are less likely to be up to date on cervical cancer screening than African American women and have the lowest participation rate in CRC screening. MENA women choose cancer screenings differently from the general US population. Instead of half obtaining both cancer screenings, only 18% are up to date for both, 50% have only cervical cancer screening, 7% CRC screening alone and 20% report no screening at all (**Harper DM 2021**)^4^.

Personal risk perceptions, and cancer perceptions and knowledge as well as physician communication behaviors, are powerful influencers of screening uptake and used to assess their influence on cancer prevention behaviors. In particular, fatalism about cancer leads to less participation in cancer screening programs among white, African American and Hispanic race/ethnicities (**(Niederdeppe 2007, Chavez LR 1997**).^5-6^ Greater perceptions of self-cancer risk relative to other people have been linked with an increase uptake of CRC screening.(**Zhao 2016, Blalock 1990, Atkinson 2015**)^7-9^

What about the modesty issues we previously found?

In addition, physician communication behaviors may be a key modifiable determinate of cancer screening behaviors (**Peterson EB 2016)**^**10**^ so much so that increasing communication behavior is included as a major objective in the *Healthy People 2030* guidelines (**HP2030)**^11^. Simple physician recommendations for a prevention intervention have been successful in increasing HPV vaccine uptake (**Kempe A, 2019**)^12^, CRC screening (**Hudson SV 2012, Atkinson 2015**)^**13-14**^ and cervical cancer screening (**Luque 2018)** ^15^. But more specific communication patterns that examine involvement in health care decisions and dealing with women’s feelings of uncertainty, might better deconstruct the association with cancer screening behaviors.

The primary aim of our study was to determine if colorectal and/or cervical cancer screening uptake among a multiethnic sample of women 50-65 years old from southeast Michigan was associated with personal risk perceptions, cancer risk perceptions and knowledge, as well as physician communication behavior.

## Methods

### Survey Measures

A cross-sectional health survey that included screening determinants questions from the NCI HINTS 5 study was developed and piloted for southeast Michigan (**Harper DM 2021**)^3^. Only women 50-65 years old who answered questions about cervical and CRC cancer screening were included in this study. Questions about hysterectomy and colectomy were not included in the survey. Prior demographic descriptors significant to this screening population were included in the analyses [**Harper DM 2021]**^4^.

The respondent’s perceptions of the communication behaviors of her primary care physician were assessed with 7-items adapted from the Consumer Assessment of Health Plans Study **(Marshall, 2001**) ^16^. These include questions such as “Did your primary care physician give you the chance to ask all the health-related questions you had?” on a four-point Likert scale (1-4): always, usually, sometimes, and never. The risk perception and knowledge section of HINTS were developed from the Health Belief Model, the Precaution Adoption Model, the Transactional Model of Stress and Coping, the Self-Regulation Model of Health Behavior and the Protection Motivation Theory (**Vernon 1999)** ^17^. The seven questions included items such as “When I think about cancer, I automatically think about death” which were also scored on a four point Likert scale (1-4): strongly disagree, somewhat disagree, somewhat agree, strongly agree. (all questions are in the **Supplemental methods**). While these two sets of seven questions are often aggregated for a summary score, we found the detailed questions provided more insight about the screening behaviors.

For the MENA sample, the survey was distributed at 12 diverse settings of MENA interest across the three southeast Michigan counties. MENA women completed the survey using online forms or paper forms, in Arabic or English, and at home or by interviewer. The remaining sample was obtained via telephone and online methods. A landline or cellphone random dial phone interview conducted by Harris Interactive Inc sampled White and African American adults. In the Rogel Cancer Center catchment area, which comprised about 2/3 of the state. An online sample was recruited through Dynata which oversampled African Americans in the catchment area. All respondents received an incentive at completion.

### Screening outcome measures

Cervical cancer screening was considered up to date if the respondent indicated a Pap and/or HPV test within the past 3 years (**USPSTF 2018)** ^18^. For colorectal cancer screening, up to date included fecal occult blood test (FOBT) or fecal immunochemistry test (FIT) within the past year or a colonoscopy within the past 10 years in accordance with the USPSTF guidelines (**USPSTF 2016)** ^19^.

### Statistical Analysis

The four screening categories were defined as those women up to date with both CRC and cervical cancer screenings, with cervical cancer screening alone, with CRC screening alone and neither screening test. The demographic data were analyzed by means and frequencies. The answers to the communication behavior questions and the risk related questions were collapsed (always/usually vs. sometimes/never) and dichotomized (strongly/somewhat disagree vs. somewhat/strongly agree) for Kruskal Wallis analysis across the four screening categories. Multinomial logistic regression modeling was used to determine the significantly different communication and risk predictors for the four screening categories adjusting for the demographic variables resulting in six different comparisons. It is important to this study that comparator screenings were more than the no screening option, which has been the most commonly reported method of analysis. It is clear from the screening rates that women screen for CRC and cervical cancer differently. Allowing the analysis to include each of these screens alone as a comparator may provide insights into associations with the differential screening outcomes. Statistical analysis was completed with Statistica v 13.2 (**Dell 2016**) ^20^.

## Results

There were 394 women, average age of 57.7 years (SD 4.5), who completed the CRC and cervical cancer screening questions. Of these, 69% self-declared as White, 19% Black and 9% MENA race (the other 3% were excluded from this analysis). The majority had a college education, were married, had a chronic disease and had an income between $50,000-$99,999. (**Table 1**).

**Table 1.** Demographic descriptors of all screening groups

|  |  |  |
| --- | --- | --- |
| **Age, yrs (mean , SD)** | 57.7 | 4.5 |
| **Race** | **N** | **%** |
| White | 271 | 68.78 |
| African American/Black | 75 | 19.04 |
| Other | 11 | 2.79 |
| MENA | 37 | 9.39 |
| **Education** |  |  |
| High School or less | 85 | 21.6 |
| Some College | 123 | 31.2 |
| College Graduate | 138 | 35.0 |
| Post college education | 48 | 12.2 |
| **Marital Status** |  |  |
| Married/partnered | 236 | 59.9 |
| Single | 156 | 39.6 |
| **Income** |  |  |
| <\$10,000 | 28 | 7.1 |
| \$10-\$49999 | 148 | 37.6 |
| \$50-\$99999 | 140 | 35.5 |
| ≥\$100,000 | 70 | 17.8 |
| **Any Chronic Condition** | 291 | 73.9 |

The four screening categories had significantly different proportions of agreement for the cancer risk perceptions and knowledge questions **(Table 2**). In particular those with neither screening had the highest agreement (strongly/somewhat) rate (33%) for the fatalism construct of ““There’s not much you can do to lower your chances of getting cancer” (p=0.03 H(3, N=387) = 9.04), whereas those who had completed both CRC and cervical cancer screening had the lowest agreement rate (15%). Likewise, “When I think about cancer, I automatically think about death” had the lowest agreement rate (48%) among those who had completed both CRC and cervical cancer screening (p=0.04 H(3,N=388)=8.22) and the highest among those who had completed only cervical cancer screening (66%).

**Table 2.**
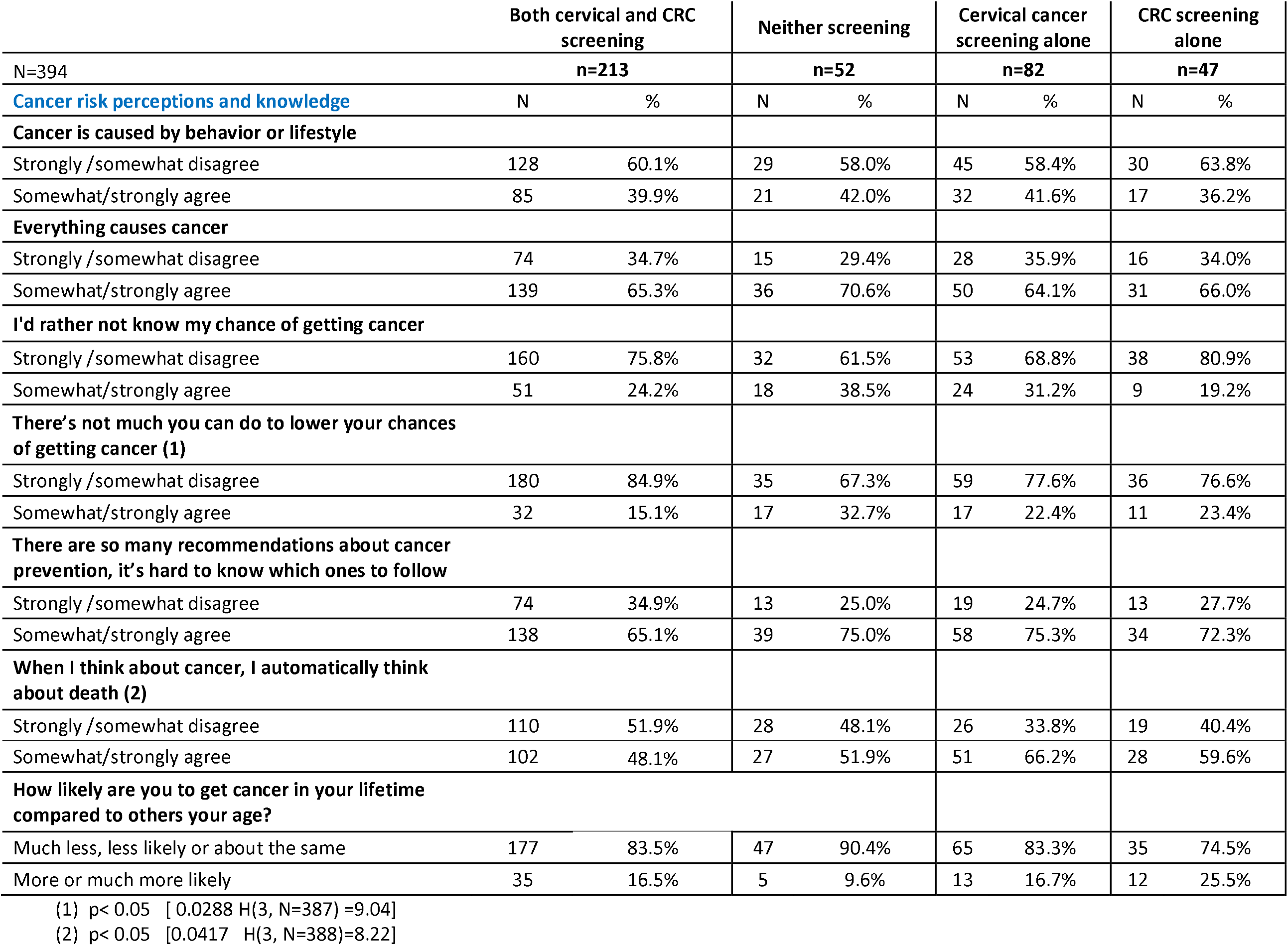
Risk perceptions and cancer knowledge by screening categories

The communication behaviors of the primary care physician was significantly different for six of the seven questions for women in each of the four screening populations (**Table 3**). The strongly/somewhat disagreement perception was lowest for those completing both CRC and cervical cancer screening in all six questions. The highest rates of disagreement were generally reported for those women who completed only a single screen. Those completing neither screen had the highest disagreement rate (24%) only when responding to “Involved you in the decisions about your health care as much as you wanted.

**Table 3.**
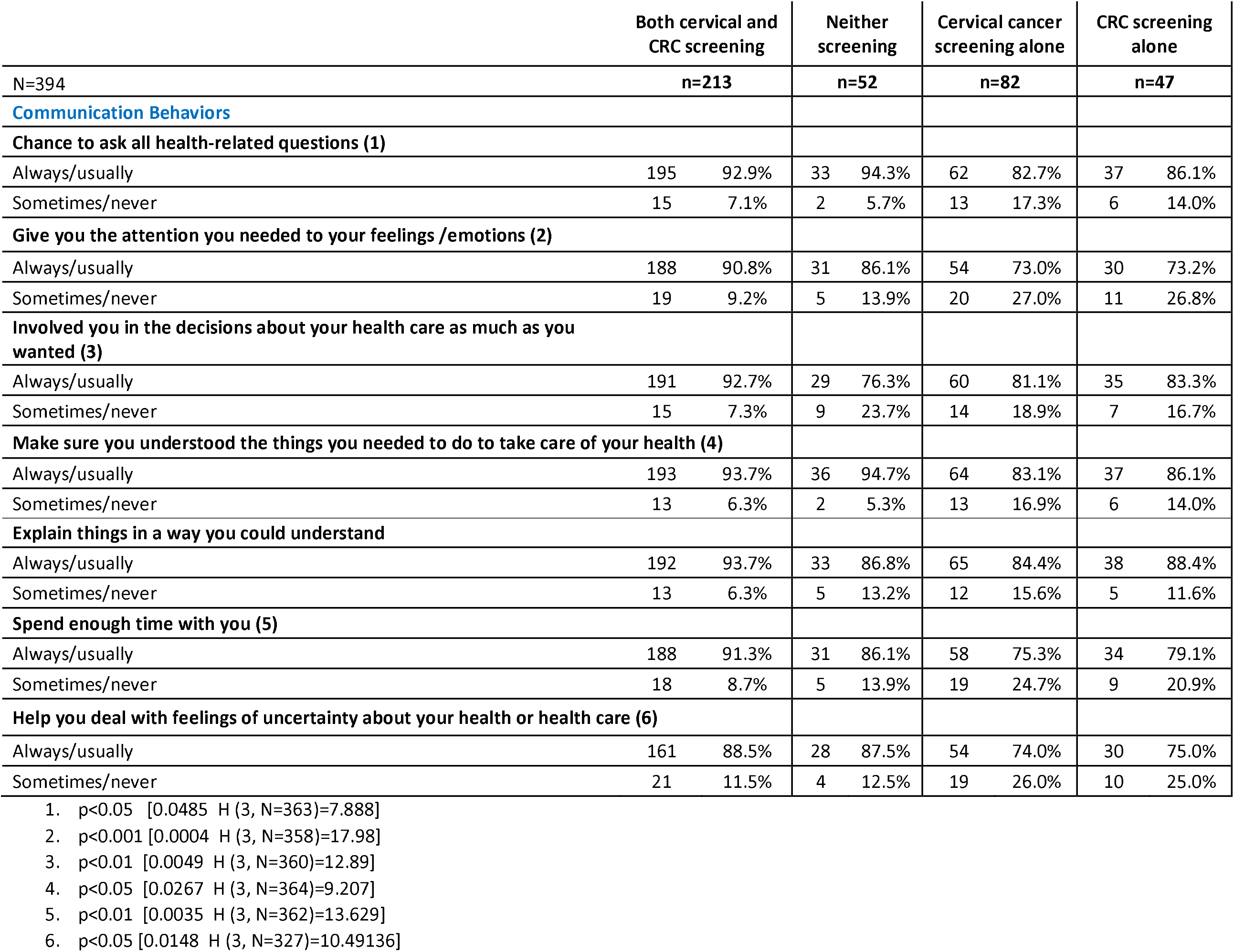
Primary care physician communication behaviors associated with the screening categories

### Multinomial modeling

All cancer risk perception statements were modeled for predicting screening behavior. **Table 4** shows that two population comparisons were influenced by different cancer risk perception questions. Those women who had completed both CRC and cervical cancer screening compared to those not doing either screen were 1.65 times more likely to somewhat/strongly disagree that “there is not much you can do to lower your chances of getting cancer” (aOR 1.65, (95% CI: 1.15, 2.35)). On the other hand, the women completing both a CRC and cervical cancer screen compared to completing only the cervical cancer screen were 41% more likely to somewhat/strongly disagree that “when they think about cancer, they automatically think about death” (aOR 1.41 (95% CI: 1.07, 2.86)).

**Table 4.** Predictors of screening by cancer risk perception and knowledge

|  | BOTH screens compared to NEITHER |  |  | BOTH compared to a cervix only |  |  |
| --- | --- | --- | --- | --- | --- | --- |
| Cancer risk perception and knowledge | aOR | L95 | U95 | aOR | L95 | U95 |
| **There's not much you can do to lower your chances of getting cancer** |  |  |  |  |  |  |
| Strongly /somewhat disagree | **1.65** | **1.16** | **2.35** | 1.18 | 0.85 | 1.66 |
| Somewhat/strongly agree | 1.00 |  |  | 1.00 |  |  |
| **When I think about cancer, I automatically think about death** |  |  |  |  |  |  |
| Strongly /somewhat disagree | 1.00 | 0.73 | 1.36 | **1.41** | **1.07** | **1.86** |
| Somewhat/strongly agree | 1.00 |  |  | 1.00 |  |  |

Bold/red is significant
No other screening population comparison was significant
No other beliefs about cancer were significant

When modeled by multivariate analyses, only one communication behavior of the six previously showing differences among screening categories remained significant, “Did your PCP involve you in the decisions about your health care as much as you wanted?” (**Table 5**) This communication behavior was significant in the following categories. Those women who had completed both CRC and cervical cancer screenings compared to those women not up to date with either screen were 99% more likely to agree that “her primary care physician involved her in the decisions about her health care as much as she wanted” (aOR=1.99, (95% CI: 1.26, 3.14)). Women who had both screens compared to only the cervical cancer screen were 72% more likely to agree that her primary care physician involved her in the decisions about her health care as much as she wanted” (aOR=1.72 (95% CI: 1.16, 2.55)).

**Table 5.** Communication behaviors predicting screening

|  | BOTH screenings<br>compared to<br>NEITHER |  |  | BOTH screenings<br>compared to Cervix<br>only |  |  |
| --- | --- | --- | --- | --- | --- | --- |
| Communication behavior | aOR | L95 | U95 | aOR | L95 | U95 |
| **Did your PCP involve you in the decisions about your health care as much as you wanted** |  |  |  |  |  |  |
| Always/usually | **1.99** | **1.26** | **3.14** | **1.72** | **1.16** | **2.55** |
| Sometimes/never | 1.0 |  |  | 1.0 |  |  |

PCP means primary care physician
Bold/red is significant
No other communication behavior was significant
No other screening population comparison was significant

When the multinomial population models were adjusted for age, race and having any chronic disease, in addition to the cancer risk perceptions and the communication behaviors, five comparisons had significant predictors (**Table 6**). None of the cancer risk perceptions were significant, nor were having any chronic diseases. The only significant predictor for women having both screens compared to neither screen was the primary care physician communication behavior to involve the woman as much as she wanted in her health care. However, the only significant predictor for women having only a CRC screening compared to no screen, was race, where MENA women were more likely than white women to have only a CRC screen than no screen. Race, likewise, was the only predictor for women who had both screens compared to only having the CRC screen where MENA and Black women were significantly less likely than white women.

**Table 6.** Communication behavior, age and race predict screening

|  | BOTH screens compared to NEITHER |  |  | CRC alone compared to NEITHER |  |  | BOTH compared to CRC alone |  |  | CRC only compared to Cervix alone |  |  | BOTH compared to Cervix alone |  |  |
| --- | --- | --- | --- | --- | --- | --- | --- | --- | --- | --- | --- | --- | --- | --- | --- |
|  | aOR | L95 | U95 | aOR | L95 | U95 | aOR | L95 | U95 | aOR | L95 | U95 | aOR | L95 | U95 |
| Age | 1.06 | 0.98 | 1.15 | 1.11 | 1 | 1.23 | 0.96 | 0.88 | 1.04 | **1.21** | **1.1** | **1.33** | **1.16** | **1.08** | **1.24** |
| RACE |  |  |  |  |  |  |  |  |  |  |  |  |  |  |  |
| White | 1 |  |  | 1 |  |  | 1 |  |  | 1 |  |  | 1 |  |  |
| Black | 1.75 | 0.76 | 4.01 |  |  |  | **0.02** | **0.01** | **0.03** |  |  |  | 1.45 | 0.73 | 2.88 |
| MENA | 0.42 | 0.17 | 1.08 | **14.8** | **1.34** | **162.8** | **0.03** | **0.00** | **0.27** | **11.61** | **1.2** | **112.05** | **0.33** | **0.15** | **0.76** |
| Did your PCP involve you in the decisions about your health care as much as you wanted? |  |  |  |  |  |  |  |  |  |  |  |  |  |  |  |
| Always/usually | **1.94** | **1.20** | **3.14** | 1.24 | 0.69 | 2.2 | 1.57 | 0.96 | 2.57 | 1.06 | 0.62 | 1.82 | **1.67** | **1.08** | **2.57** |
| Sometimes/never | 1 |  |  | 1 |  |  | 1 |  |  | 1 |  |  | 1 |  |  |

PCP means primary care physician
Bold/red is significant
Greyed out means insufficient numbers for stable analysis

There were two predictors for women having only the CRC screen compared to having only the cervical cancer screen: age and race. The older the woman, the more likely she would have only a CRC screen and not a cervical cancer screen. Likewise, MENA women compared to white women were more likely to have only a CRC screen compared to only a cervical cancer screen.

Finally, three distinct predictors informed who had both CRC and cervical cancer screenings compared to only having cervical cancer screening. These were older age, MENA vs. white race and the physician’s communication behavior to involve her in her health care as much as she wanted.

## Discussion

We focused on understanding the many combinations of screening uptake that occur and found different predictors for CRC and cervical cancer screening together or alone. In prior work, having a chronic disease increased CRC and/or cervical cancer screening rates compared to no screen (**Harper 2021**,**Cofie 2019, Borrayo 2016**)^3, 21-22^and other studies have shown that cancer fatalism is a major barrier to screening uptake (**Clarke 2021, Jun 2013**).^23-24^ In this work, neither having chronic diseases nor cancer risk perceptions, such as cancer fatalism, had an influence on any cancer screening behavior pattern. We did however find that positive provider communication that includes the woman in her health care as much as she wants was associated with completing both CRC and cervical cancer screening compared to no screen, but even more importantly compared to only cervical cancer screening- **the first new insight from this work**. These results may indicate that a single screen when done outside the medical home is perceived as less care for the whole person. We know some primary care physicians do not include pelvic exams in their scope of practice, whereas other programs have implemented targeted screening to increase cervical cancer screening as an endpoint in itself [**Wong 2019, Pollack 2020**].^25-26^ While these practice patterns and programs may provide a pathway to reach a single cancer screening goal, they may be detrimental to the whole patient care where the patient physician dyad works as a team to provide options for best health. Our work emphasizes the importance of the physician involving the woman as much as she wants in her own health care as a powerful facilitator associated with both CRC and cervical cancer screenings. We hypothesize that this relationship may further enhance follow up after any abnormal screenings (**Tsui 2019, Peterson 2016**).^27-28^

Age continues to be an important predictor of these two screens. Within the 50-65 year age range, younger women participate more in the cervical cancer screen and older women participate more in the CRC screen. **This division is the second new insight from this work**. We do not know the causes of the age divide by cancer screening. Future work is needed to examine ways to overcome this divide.

Moreover, the MENA women were important to include in this work because health care disparities in the MENA community are not readily known due to how MENA status is obtained in the US Census. Currently it must be indicated by having the respondent write in their MENA heritage rather than having it as a separate preidentified racial category. We have shown in this work that MENA women are rarely screened for CRC cancer, be it alone or in addition to cervical cancer screening. Past work shows little CRC screening alone (**Harper 2021**) ^3^. Targets for improvement of MENA women’s health including cancer screens need to encompass physician communication training, and patient education about the options for CRC screening as part of whole person health care.

Most importantly this work shows that race, age and physician communication behavior are three independent influencers of completing both CRC and cervical cancer screening compared to cervical cancer screening alone. Future research will focus on a qualitative study asking women why they complete one screen and not the other.

## Strengths and Limitations

The strengths of this study include comparing differential screenings for cervical and CRC whereas all prior literature has only compared screening to no screening. In addition these two specific organ screenings were intentionally paired in a novel way because their barriers are quite similar: “Among racially diverse populations with less than a high school education, low income, no health insurance, and no regular health care provider, other barriers to CRC screening are fatalism, religious beliefs, lack of self-worth, sexually related concerns, history of sexual abuse, and past negative experiences with screening” (**Knight JR 2015**) ^29^. These are the same barriers that others have shown for cervical cancer screening **(Harper 2020)** ^1^.

Understanding the differences for completion of one screen but not another is important to understand as prior screening strategies targeting a single screen may have missed the opportunity to present options for other cancer screenings.

Another strength of this study is the inclusion of MENA women who have little representation in the literature. We have been able to show new deficits in CRC screening not previously identified.

Moreover, our methodologic approach is a strength in that epidemiologic risk factors for more than one cancer screen are not often accompanied in the analysis by communication and risk perception understanding.

The limitations of this study, on the other hand, include the cross-sectional survey design. It is not possible to infer causal relationships between constructs or items in the survey. In addition, while the sample size is adequate for our statistical analyses, there were cells sizes with insufficient numbers for all models to converge or have generalizable results. Moreover, all outcomes were self-reported with an opportunity to over-report actual screening frequencies (**Bonafede 2019**).^30^

This survey was administered with three approaches, where the online and random digit dialing were methods not used with the MENA population. Despite this, the population weighting previously done (**Harper 2021**)^3^ provides full generalizability to Southeast Michigan.

## Conclusions

We have shown that CRC and cervical cancer screening, with similar barriers, are not binary activities. There are different reasons that women partake in one screen and not the other. These reasons include age, race and relationship with the physician. We can no longer separate screening for these two anatomically proximate cancers from each other.

## Data Availability

This is the Rogel Cancer Center Catchment Survey, funded by the NCI, and will be made available to interested parties.

## References

1. Harper DM, Plegue M, Harmes KM, Jimbo M, SheinfeldGorin S. Three large scale surveys highlight the complexity of cervical cancer under-screening among women 45-65 years of age in the United States. Prev Med, 2020;130:105880. doi: 10.1016/j.ypmed.2019.105880

2. American Cancer Society. Colorectal Cancer Facts & Figures 2020-2022. Atlanta: American Cancer Society; 2020 https://www.cancer.org/research/cancer-facts-statistics/colorectal-cancer-facts-figures.html

3. Harper DM, Plegue M, Sen A, Gorin SS, Jimbo M, Patel MR, Resnicow K. Predictors of screening below the belt for cervical and colorectal cancer in women 50-65 years old in a multi-ethnic population. Prev Med Rep, 2021; 22:101375. doi.org/10.1016/j.pmedr.2021.101375

4. Harper DM, Tariq M, Alhawli A, Syed N, Patel M, Resnicow K. Comparative predictors for cervical cancer screening by population group: Middle Eastern-North African (MENA), White and African American/Black women. JABFM, 2021

5. Niederdeppe J, Levy AG. Fatalistic beliefs about cancer prevention and three prevention behaviors. Cancer Epidemiol Biomarkers Prev, 2007 May;16(5):998–1003. doi: 10.1158/1055-9965.EPI-06-0608.

6. Chavez LR, Hubbell FA, Mishra SI, Valdez RB. The influence of fatalism on self-reported use of Papanicolaou smears. Am J Prev Med, 1997;13(6):418–24. PMID: 9415785 https://doi.org/10.1016/S0749-3797(18)30134-X

7. Zhao X, Nan X. The influence of absolute and comparative risk perceptions on cervical cancer screening and the mediating role of cancer worry. J Health Commun, 2016;21(1):100–8. doi: 10.1080/10810730.2015.1033114.

1. Blalock, SJ, DeVellis BM, Afifi RA, Sandler RS. Risk perceptions and participation in colorectal cancer screening. Health Psychology, 1990;9:792–806. doi:10.1037=0278-6133.9.6.792

2. Atkinson TM, Salz T, Touza KK, Li Y, Hay JL. Does colorectal cancer risk perception predict screening behavior? A systematic review and meta-analysis. J Behav Med, 2015;38(6):837–50. doi: 10.1007/s10865-015-9668-8.

3. Peterson EB, Ostroff JS, DuHamel KN, D’Agostino TA, Hernandez M, Canzona MR, Bylund CL. Impact of provider-patient communication on cancer screening adherence: A systematic review. Prev Med, 2016;93:96–105. doi: 10.1016/j.ypmed.2016.09.034

4. HP 2030 Guidelines.https://health.gov/healthypeople/objectives-and-data/browse-objectives/cancer/increase-proportion-people-who-discuss-interventions-prevent-cancer-their-providers-c-r02

5. Kempe A, O’Leary ST, Markowitz LE, Crane LA, Hurley LP, Brtnikova M, Beaty BL, Meites E, Stokley S, Lindley MC. HPV vaccine delivery practices by primary care physicians. Pediatrics, 2019;144(4):e20191475. doi: 10.1542/peds.2019-1475.

6. Hudson SV, Ferrante JM, Ohman-Strickland P, Hahn KA, Shaw EK, Hemler J, Crabtree BF. Physician recommendation and patient adherence for colorectal cancer screening. J Am Board Fam Med, 2012;25(6):782–91. doi: 10.3122/jabfm.2012.06.110254.

7. Atkinson TM, Salz T, Touza KK, Li Y, Hay JL. Does colorectal cancer risk perception predict screening behavior? A systematic review and meta-analysis. J Behav Med, 2015;38(6):837–50. doi: 10.1007/s10865-015-9668-8.

8. Luque JS, Tarasenko YN, Chen C. Correlates of cervical cancer screening adherence among women in the U.S.: Findings from HINTS 2013-2014 J Prim Prev, 2018;39(4):329–44. doi: 10.1007/s10935-018-0513-z.

9. Marshall GN, Morales LS, Elliot M, Spritzer K, Hays RD. Confirmatory factor analysis of the consumer assessment of health plans study (CAHPS) 1.0 Core Survey. Psychol Assess, 2001:13(2):216–29. doi: 10.1037//1040-3590.13.2.216.

10. Vernon SW. Risk perception and risk communication for cancer screening behaviors: A review. J Natl Cancer Inst Monogr, 1999;(25):101–19. doi:10.1093/oxfordjournals.jncimonographs.a024184

11. US Preventive Services Task Force, Curry SJ, Krist AH, Owens DK, Barry MJ, Caughey AB, Davidson KW, Doubeni CA, Epling JW Jr, Kemper AR, Kubik M, Landefeld CS, Mangione CM, Phipps MG, Silverstein M, Simon MA, Tseng CW, Wong JB. Screening for cervical cancer: US preventive services task force recommendation statement. JAMA, 2018;320(7):674–86. doi: 10.1001/jama.2018.10897.

12. US Preventive Services Task Force, Bibbins-Domingo K, Grossman DC, Curry SJ, Davidson KW, Epling JW Jr, García Far, Gillman MW, Harper DM, Kemper AR, Krist AH, Kurth AE, Landefeld CS, Mangione CM, Owens DK, Phillips WR, Phipps MG, Pignone MP, Siu AL. Screening for colorectal cancer: US preventive services task force recommendation statement. JAMA, 2016;315(23):2564–75. doi: 10.1001/jama.2016.5989

13. Dell Inc (2016) Dell Statistica (data analysis software system), version 13.2. software.dell.com.

14. Cofie LE, Hirth JM, Wong R. Chronic comorbidities and cervical cancer screening and adherence among US-born and foreign-born women. Cancer Causes Control, 2018;29(11):1105–13. doi: 10.1007/s10552-018-1084-2.

15. Borrayo BD, O’Lawrence H. A post analysis of a preventive and chronic healthcare tool. J Health Hum Serv Adm, 2016;39(1):15–40. PMID: 27483973

16. Clarke N, Kearney PM, Gallagher P, McNamara D, O’Morain CA, Sharp L. Negative emotions and cancer fatalism are independently associated with uptake of faecal immunochemical test-based colorectal cancer screening: Results from a population-based study. Prev Med, 2021;145:106430. doi: 10.1016/j.ypmed.2021.106430.

17. Jun J, Oh K. 2013 Asian and Hispanic Americans’ cancer fatalism and colon cancer screening. Am J Health Behav, 2013;37(2):145–54. doi: 10.5993/AJHB.37.2.1.

18. Wong FL, Miller JW. Centers for Disease Control and Prevention’s National Breast and Cervical Cancer Early Detection Program: Increasing access to screening. J Womens Health (Larchmt), 2019;28(4):427–31. doi: 10.1089/jwh.2019.7726.

19. Pollack LM, Ekwueme DU, Hung MC, Miller JW, Chang SH. Estimating the impact of increasing cervical cancer screening in the National Breast and Cervical Cancer Early Detection Program among low-income women in the USA. Cancer Causes Control, 2020;31(7):691–702. doi: 10.1007/s10552-020-01314-z.

20. Tsui J, Llanos AA, Doose M, Rotter D, Stroup A. Determinants of abnormal cervical cancer screening follow-up and invasive cervical cancer among uninsured and underinsured women in New Jersey. J Health Care Poor Underserved, 2019;30(2):680–701. doi: 10.1353/hpu.2019.0050.

21. Peterson EB, Ostroff JS, DuHamel KN, D’Agostino TA, Hernandez M, Canzona MR, Bylund CL. Impact of provider-patient communication on cancer screening adherence: A systematic review. Prev Med, 2016;93:96–105. doi: 10.1016/j.ypmed.2016.09.034.

22. Knight JR, Kanotra S, Siameh S, Jones J, Thompson B, Thomas-Cox S. Understanding barriers to colorectal cancer screening in Kentucky. Prev Chronic Dis, 2015;12:E95. doi: 10.5888/pcd12.140586.

23. Bonafede MM, Miller JD, Pohlman SK, Troeger KA, Sprague BL, Herschorn SD, Winer IH. Breast, cervical, and colorectal cancer screening: Patterns among women with medicaid and commercial insurance. Am J Prev Med, 2019;57(3):394–402. doi: 10.1016/j.amepre.2019.04.010.

