## Supplemental Table 1 for "Cancer risk perception and knowledge and physician communication behaviors: specific influences on cervical cancer and colorectal cancer screening in women 50-65 years old"

Supplement Methods Questions

**Cancer risk perception and knowledge**

**Caused by behavior or lifestyle**

**Everything causes cancer**

**I'd rather not know my chance of getting cancer**

**There's not much you can do to lower your chances of getting cancer***

**There are so many recommendations about cancer prevention, it’s hard to know which ones to follow**

**When I think about cancer, I automatically think about death***

**How likely are you to get cancer in your lifetime compared to others your age?**

**Physician communication behavior**

**Chance to ask all health-related questions**

**Give you the attention you needed to your feelings /emotions**§

**Involved you in the decisions about your health care as much as you wanted Ɨ**

**Make sure you understood the things you needed to do to take care of your health**

**Explain things in a way you could understand**

**Spend enough time with you**

**Help you deal with feelings of uncertainty about your health or health care**
